# A curriculum-based approach to make healthcare inclusive for the transgender population: challenges and lessons from Pakistan

**DOI:** 10.1101/2023.09.26.23295951

**Authors:** Akash Kumar Ahuja, Manzar Abbas, Mian Arsam Haroon, Muhammad Abdullah Javed, Areeba Memon, Rida Irfan, Zohair Karim, Areesh Bhatti, Mehsa Hashim, Janeeta Hamid, Asaad Nafees

## Abstract

**Background:** Transgender people experience significant healthcare inequalities due to stigma and lack of acceptance. Physicians and medical students have reported knowledge gaps regarding transgender health care (TGHC). Therefore, we conducted this study to assess the perceived need for and preferred approaches of medical students towards incorporation of TGHC in curriculum and any possible barriers that can arise.

**Methods:** A cross-sectional survey was conducted amongst medical students from accredited medical colleges in Pakistan. Google forms were used to collect data including three outcome variables: need, preferred modalities, and barriers towards TGHC. The outcome variables were analyzed and compared using SPSS version 25.0.

**Results:** A total of 436 medical students from all over the Pakistan responded to the questionnaire. Of those, 264 (60.6%) were female. A majority of participants were from the southern provinces of Pakistan (n = 302; 69.3%). An overwhelming majority (98.4 %) of participants (n=429) felt there was a need to teach transgender healthcare (TGHC) education at the undergraduate level. This is contrasted by only 5% of participants (n=23) reporting being taught transgender health as part of their curriculum. Of these 5%, knowledge was mostly assessed either on multiple choice or essay questions. Majority students reported never having had any interaction with transgender patient during their clinical rotations. 82.8% students identified that education provided at their institute regarding transgender health education is inadequate while 80.8% students revealed that transgender health education was not an official part of their institute’s curriculum. The biggest barrier identified was TGHC not being identified as a problem, followed by cultural and logistic barriers. The most recommended mode of TGHC education delivery as suggested by students was the use of small group discussions followed by student presentations and use of films and documentaries.

**Conclusion:** A clear gap exists regarding transgender health education in the medical curriculum in Pakistan. The study highlights important barriers and recommends possible modes of delivery to inculcate TGHC education in the curriculum.

## Introduction

The term “transgender” refers to individuals who identify as a sex other than that which was assigned to them at birth. Transgender people numbering about 25 million worldwide, experience significant healthcare inequalities (1). They are also frequently the target of sexual and gender-based violence and discrimination (2).

Transgender people continue to face significant barriers when it comes to receiving the health care they seek, due to stigma and lack of acceptance (3,4). Healthcare systems are grossly unable to meet their demands, with the main impediment being a scarcity of practitioners trained in trans-health. Due to inadequate education and exposure, physicians and medical students report knowledge gaps.

Transgender individuals are at an increased risk for mental health disorders, cancer and substance abuse (5). They are at a higher risk of sexually transmitted illnesses like HIV due to their reduced participation in health promotion and disease prevention activities, particularly those linked to sexual health. With a 49-fold higher risk of infection compared to the general population, they are one of the most vulnerable populations in the HIV epidemic (6). Moreover, Shinde et al. Reported prevalence of 32% for sexually transmitted infections such as syphilis, gonorrhea and chlamydia in transgender population with a majority of them reporting to have genital scabies and warts around perianal region (7). The situation is more pronounced in low- and middle-income countries (LMICs) such as Pakistan where the HIV incidence amongst transgender people contributes to 17°5% of the entire HIV population (8).

In Pakistan, transgender individuals are underrepresented in the national census due to fear of discrimination. They are even more vulnerable to mental health issues due to the violence and social exploitation they experience (9). With the true numbers not being reported, there is a scarcity of data on transgender healthcare (TGHC) and the difficulties they encounter. Their sexuality and gender identification might have an impact on their medical rights and care. A census carried out in 2017 reported 10,418 transgender people nationally. The census recognized transgender people based on their national identity cards but did not account for those whose cards do not reveal their gender to prevent hostility. This may be attributed to financial constraints and lack of government assistance (10). Despite the legal recognition, access to quality health care is alarmingly scarce compared to their cis counterparts.

Literature suggests several interventions to educate medical students regarding trans-health (3,11). A prior study found that several pedagogical interventions, including online modules, quizzes, seminars, and interactive patient-provider panels, improved students’ gender-affirming medical skills and understanding (12). Similarly, a team-based interprofessional simulation activity was carried out over a period of two years that showed an improvement in students’ understanding of transgender healthcare (13). Additional approaches included didactic lectures (14), small group discussions followed by multimedia presentations (15–17) and social events such as film and documentary screenings or educational games. Other methods less frequently used were role play (18), panel sessions, pre-reading of study materials (19) and seminars (16). It is evident that little to no data is available on the preferred methods and educational modality of the students themselves. With such a large percentage of the population identifying as transgender, it is critical to create a society where trans individuals have the same access to healthcare as everyone else. Gender bias in routine clinical care encounters could be eliminated by including such pedagogies into the academic curriculum (20). As medical students are the doctors of tomorrow, it is crucial that they are well-equipped with knowledge and education that is sensitive to transgender people’s needs.

The rights of transgender individuals are repeatedly overlooked within societies across the globe. Differences in physiology lead to specific healthcare requirements but unfortunately medical students in lower middle-income countries (LMICs) are not well trained to appreciate and respond to these apparent differences (4). Education centers in developing countries differ from those in the developed countries in terms of resources, social and cultural norms, and the level of understanding of medical students. Tools of learning that are appreciated amongst students of developed countries might not be preferred by students of LMICs, so, further studies were needed to evaluate the best modality available. Therefore, we conducted this study to assess the needs and preferred approaches of medical students in Pakistan on incorporating TGHC education into the curriculum, and to identify the possible barriers for incorporating TGHC education into the curriculum.

## Materials and method

### Study design and setting

We conducted a cross sectional survey from 11th October 2021 to 5th November 2021. Data were collected after getting the required approval from the Ethical Review Committee at the Aga Khan University (ERC number: 2021-6814-19491). The study recruited students online across different medical colleges in Pakistan using social media.

### Study population

All medical students enrolled in accredited medical colleges of Pakistan at the time of data collection were included, while medical students enrolled in foreign medical universities, undergraduate students enrolled in programs other than MBBS, medical graduates, practicing physicians and other healthcare professionals were excluded.

### Data collection

The Google form was disseminated among the target population of the study using various social media platforms like Facebook and WhatsApp groups for medical students. Prior to starting the survey, the participants were provided with an informed consent explaining the nature of our study and their role in it.

We used a structured and validated questionnaire developed by Joanne Rolls et al. (21), with a few questions modified based on characteristics of the target population. The first section collected information about participants’ demographics, needs, perceptions, background knowledge, and preferred modes of delivery of content related to TGHC. The second section determined perceptions of participants regarding appropriate modes of assessment for content related TGHC. The last section assessed perceived barriers in the process of adding TGHC education into the medical curriculum.

### Statistical Analysis

Data were analyzed using SPSS version 25.0. The quantitative variables such as age and total number of perceived barriers were represented as mean and standard deviation. These were compared using the independent sample Student’s t-test. The qualitative variables such as association between institution type and perceived importance of TGHC education were compared using the Chi-Square test. For inferential statistics, a p-value of less than 0.05 was considered significant in our analysis.

## Results

A total of 436 medical students from all over the Pakistan responded to the questionnaire. Of these, 168 (38.5%) were male. The largest percentage of representatives were from the southern provinces of Pakistan (n = 302; 69.3%). The demographics of the respondents are shown in Table 1.

**Table 1.** Sociodemographic Characteristics (n=436)

| **Table 1. Sociodemographic Characteristics (n=436)** |  |
| --- | --- |
| **Characteristics** | **n (%)** |
| **Students** |  |
| **Gender** |  |
| Male | 168 (38.5) |
| Female | 264 (60.6) |
| Prefer not to say | 4 (0.9) |
| **Age, Mean (SD)** | 21.7 (1.70) |
| **Year of Study** |  |
| Basic Sciences * | 150 (34.4) |
| Clinical Sciences ** | 286 (65.6) |
| **Institutions** |  |
| **Type** |  |
| Public | 97 (22.2) |
| Private | 331 (75.9) |
| Military | 8 (1.8) |
| **Geographical Location** |  |
| North Pakistan *** | 134 (30.7) |
| South Pakistan **** | 302 (69.3) |
| *Years of instruction I and II<br>**Years of instruction III, IV, and V<br>***Punjab, KPK, AJK<br>****Sindh, Baluchistan |  |

### Needs and Perceptions

Of the 436 medical students, 5.3% (n=6) reported being taught some form of transgender healthcare education at their institute. 2.5% of respondents (n=11) said the annual duration of such education lasted from 1-3 hours, 2.1% of respondents (n=9) said the duration was 4-6 hours, and 0.7% of respondents (n=3) said the duration was 7-10 hours (Figure 1). 7.1% of respondents (n=31) mentioned they encountered a transgender person in their hospital inpatient or outpatient departments.

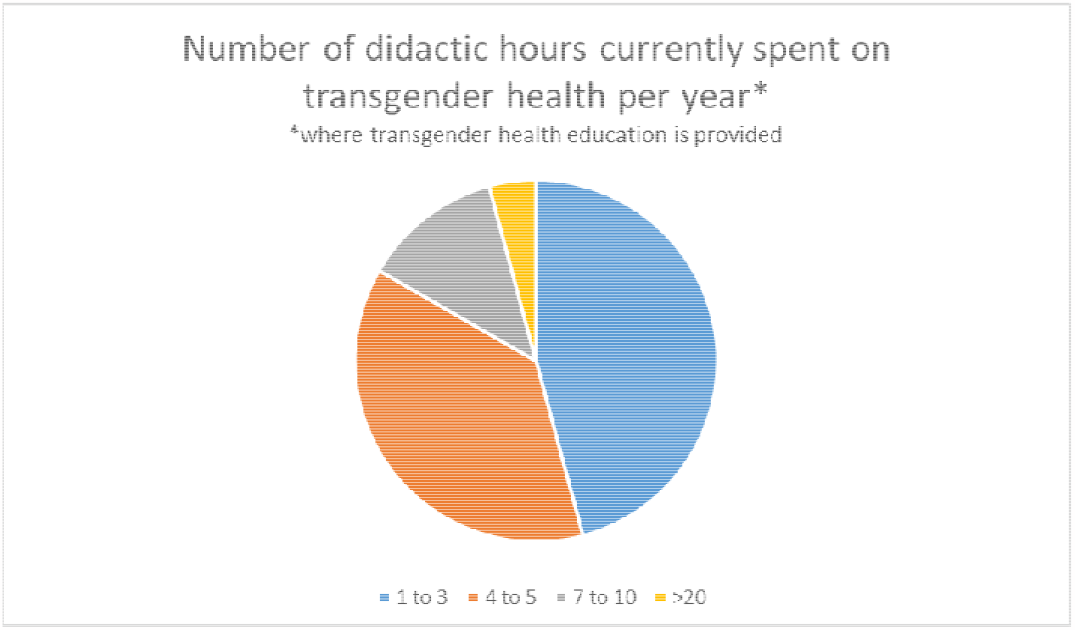

All participants responded that they felt it was important to teach transgender healthcare education at their institution. With respect to transgender healthcare coverage, 1.8% (n=8) responded it was very good, 6.2% (n=27) responded it was good, 9.4% (n=41) responded it was adequate, 33.3% (n=145) responded it was inadequate, 49.3% (n=215) responded it was poor. 9.2% (n=40) respondents reported that their institute has faculty who is knowledgeable about transgender health whereas 28.9% (n=126) responded there is no such faculty member present. The majority responded there is a need for transgender healthcare education with 98.4% (n=429) participants agreeing and 1.6% (n=7) responding with not needed. The duration of suggested hours spent on teaching transgender health varied across participants, with 2.1% (n=9) respondents reporting 0 hours to 18.1% (n=79) reporting 7-10 hours, as represented in Figure 2.

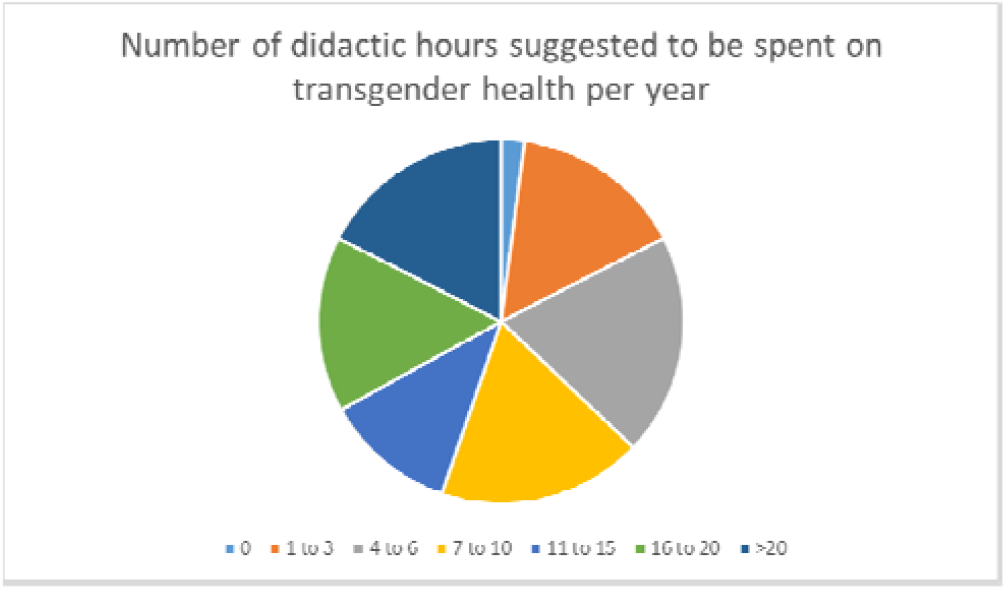

### Modes and Areas of Delivery

Only 5.3% medical students reported that their institution teaches about transgender health. The students identified the following methods as their current mode of delivery of transgender health education at their institutions include longitudinal themes sessions (10.5%), pre-reading of study materials and didactic lectures (13.2%), and seminars (15.8%). As shown in table 2, the majority of responses (18.1%) suggested that problem-based learning should be the mode of instruction to cover transgender health, followed by small group discussions (15.9%).

**Table 2.**
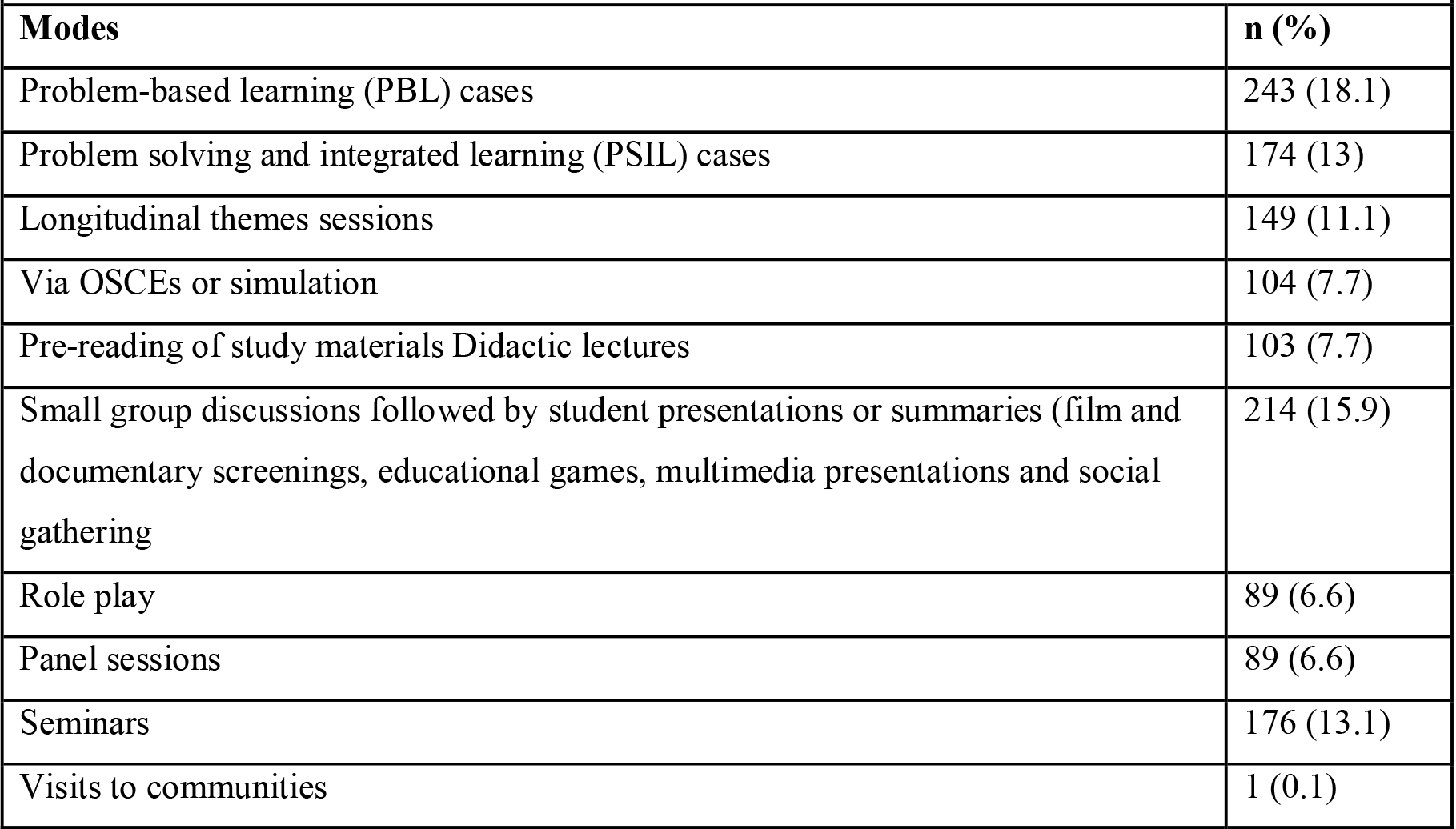
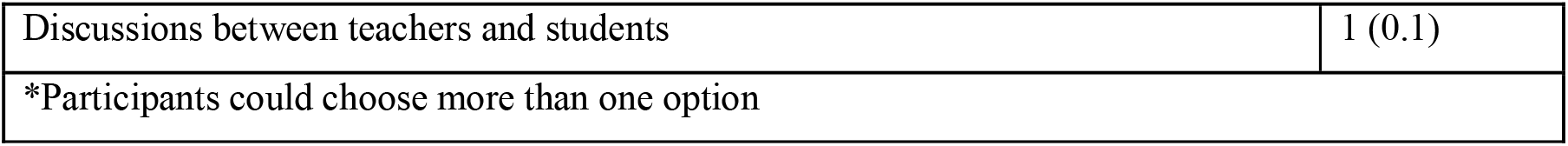
Suggested modes of transgender healthcare education (n=1343*)

| Modes | n (%) |
| --- | --- |
| Problem-based learning (PBL) cases | 243 (18.1) |
| Problem solving and integrated learning (PSIL) cases | 174 (13) |
| Longitudinal themes sessions | 149 (11.1) |
| Via OSCEs or simulation | 104 (7.7) |
| Pre-reading of study materials Didactic lectures | 103 (7.7) |
| Small group discussions followed by student presentations or summaries (film and documentary screenings, educational games, multimedia presentations and social gathering | 214 (15.9) |
| Role play | 89 (6.6) |
| Panel sessions | 89 (6.6) |
| Seminars | 176 (13.1) |
| Visits to communities | 1 (0.1) |
| Discussions between teachers and students | 1 (0.1) |
| *Participants could choose more than one option |  |

The topics that they were taught regarding transgender health included health access issues/barriers to care faced by transgender persons (14.4%), health disparities faced by transgender persons (13.7%), mental health among transgender persons (11.5%) and some others. These topics were covered in family medicine, endocrinology, woman’s health/OBGYN, urology and infectious disease, with the highest coverage in family medicine (29.7%) and endocrinology (29.7%). Some of the topics that the students suggested that they believed should be taught about transgender health are presented in table 3.

**Table 3.**
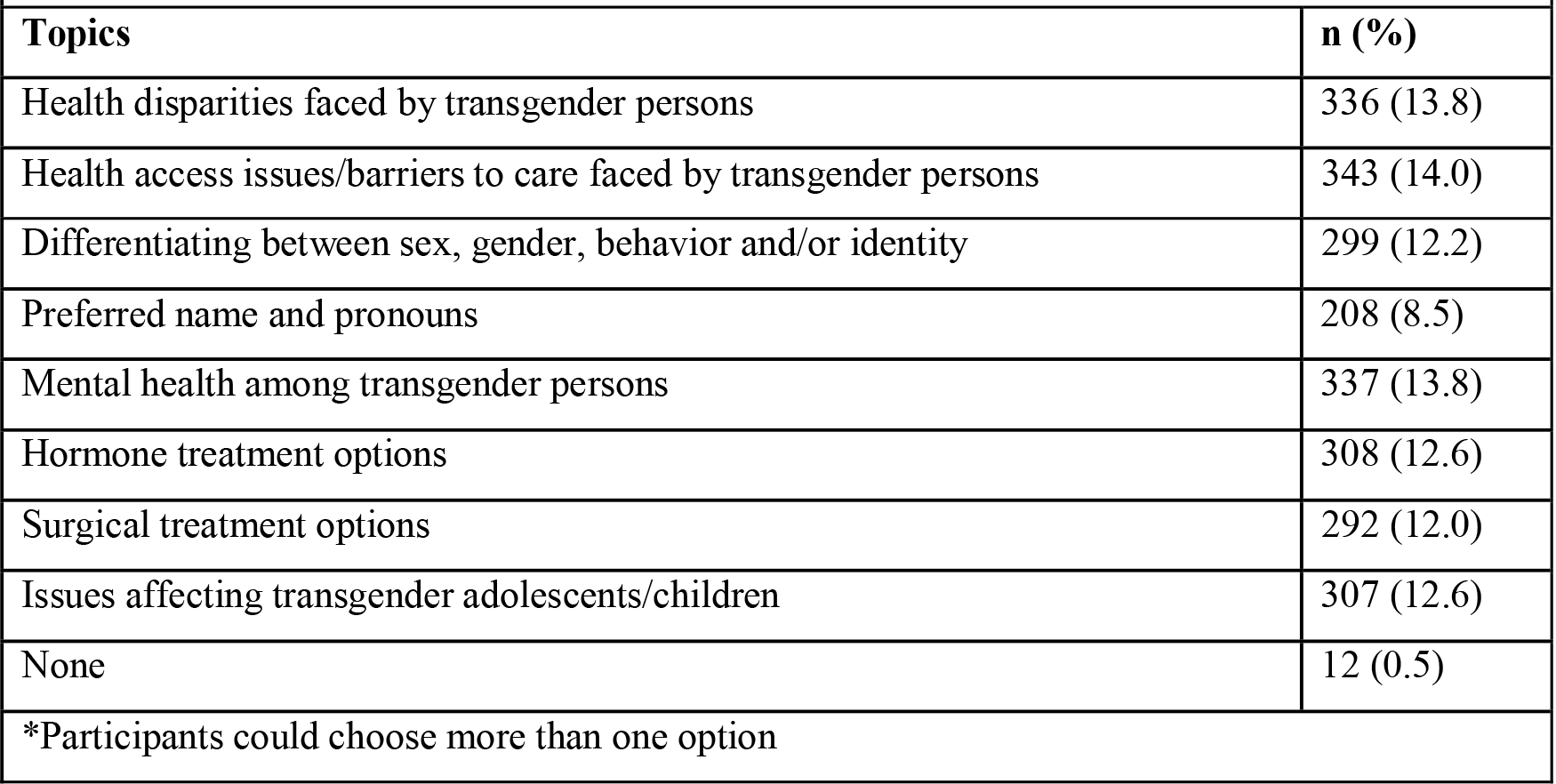
Suggested topics for instruction (n=2442 responses*)

| **Table 3. Suggested topics for instruction (n=2442 responses\*)** |  |
| --- | --- |
| **Topics** | **n (%)** |
| Health disparities faced by transgender persons | 336 (13.8) |
| Health access issues/barriers to care faced by transgender persons | 343 (14.0) |
| Differentiating between sex, gender, behavior and/or identity | 299 (12.2) |
| Preferred name and pronouns | 208 (8.5) |
| Mental health among transgender persons | 337 (13.8) |
| Hormone treatment options | 308 (12.6) |
| Surgical treatment options | 292 (12.0) |
| Issues affecting transgender adolescents/children | 307 (12.6) |
| None | 12 (0.5) |
| *Participants could choose more than one option |  |

314 (72.0%) students believed that family medicine was the most appropriate clinical rotation where these topics should be addressed followed by endocrinology (56.2%) and woman’s health/ OBGYN (48.2%).

### Perceived Barriers

The most highly reported perceived barrier to providing TGHC education at one’s institute was TGHC not considered to be an important issue (n=279, 64% of participants). This was followed by other barriers reported across the board, as shown in Table 8. It is important to note that 9.6% (n=42 students) respondents reported that no barriers existed in providing TGHC education to medical students at their programs.

### Independent Variable Analysis

We explored the association of institution type with the different variables assessed in the questionnaire using the chi square test (Table 6). We noted that whether the institute was of a public, private, or military type had no significant association with the occurrence of an interaction with a transgender patient, the instruction of transgender healthcare education, the perceived importance including TGHC in medical education, and the overall coverage of TGHC issues. There was also no association with the likelihood of a participant attending a TGHC education session (p=0.119) although an association with faculty informed about TGHC exists (p=0.005). However, our analysis indicates a significant association between institution type and the likelihood of a colleague attending a session (p=0.028). Despite no significant association existing between perceived importance and institution type, a significant relationship between the need for TGHC education and institution type was observed (p=0.006)

**Table 4.** Suggested mode of assessment (n=839 responses*)

| **Table 4. Suggested mode of assessment (n=839 responses\*)** |  |
| --- | --- |
| **Mode** | **n (%)** |
| Essay questions on an exam | 105 (12.5) |
| Multiple choice questions on an exam | 197 (23.5) |
| OSCEs | 155 (18.5) |
| Reflection | 148 (17.6) |
| Project or Presentation | 228 (27.2) |
| Should not be assessed | 6 (0.7) |
| *Participants could choose more than one option |  |

**Table 5.** Barriers in providing education about transgender health (n=1618*)

|  |  |
| --- | --- |
| **Barriers** | **n (%)** |
| Lack of time/hours in curriculum | 214 (13.2) |
| Lack of faculty knowledge about transgender health | 222 (13.7) |
| Lack of faculty comfort regarding teaching transgender health | 207 (12.8) |
| Lack of student interest/comfort | 162 (10.0) |
| Transgender health not considered to be an important issue | 279 (17.2) |
| Institutional culture not supportive of teaching about transgender health at my university | 144 (8.9) |
| Societal culture not supportive of teaching about transgender health at my university | 203 (12.5) |
| It is unclear where in the curriculum to include transgender health topics | 140 (8.7) |
| No barriers | 42 (2.6) |
| Other | 6 (0.4) |
| *Participants could choose more than one option |  |

**Table 6.** Association of institution type with variables.

| **Table 6. Association of institution type with variables** |  |  |  |
| --- | --- | --- | --- |
| **Variable** | **$\chi^2$ value** | **Degree of freedom** | **p-value** |
| Transgender patient encounter | 5.078 | 4 | 0.279 |
| Provision of TGHC education | 1.319 | 4 | 0.858 |
| Perceived Importance | 9.308 | 6 | 0.157 |
| Faculty knowledgeable about transgender health | 18.535 | 6 | 0.005 |
| Likelihood participant will attend session | 12.811 | 8 | 0.119 |
| Likelihood colleague will attend session | 17.181 | 8 | 0.028 |
| Suggested number of didactic TGHC education hours | 11.194 | 12 | 0.512 |
| Need for TGHC education | 14.604 | 4 | 0.006 |
| Assessment of TGHC education | 3.292 | 4 | 0.510 |
| Overall coverage of TGHC issues | 4.217 | 8 | 0.837 |

Our analysis in Table 7 observed multiple significant associations between institution region (i.e., North Pakistan and South Pakistan) and the variables assessed. Similar to the analysis involving institution type, a significant association was observed of TGHC knowledgeable faculty and institution region (p<0.001). Another similarity was that of the relationship of the likelihoods of attending a TGHC education session, with only that of a colleague having a significant association with institution region (p=0.001). Conversely, institution region was significantly associated with the instruction of TGHC education (p=0.020) but a relationship with the need for TGHC education was not observed (p=0.337).

**Table 7.**
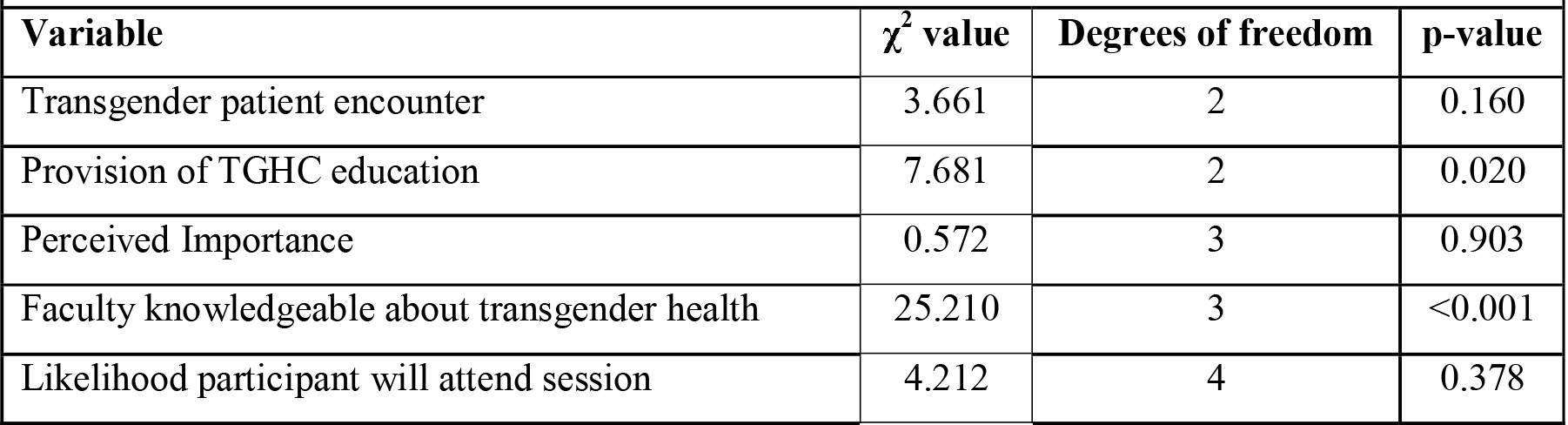
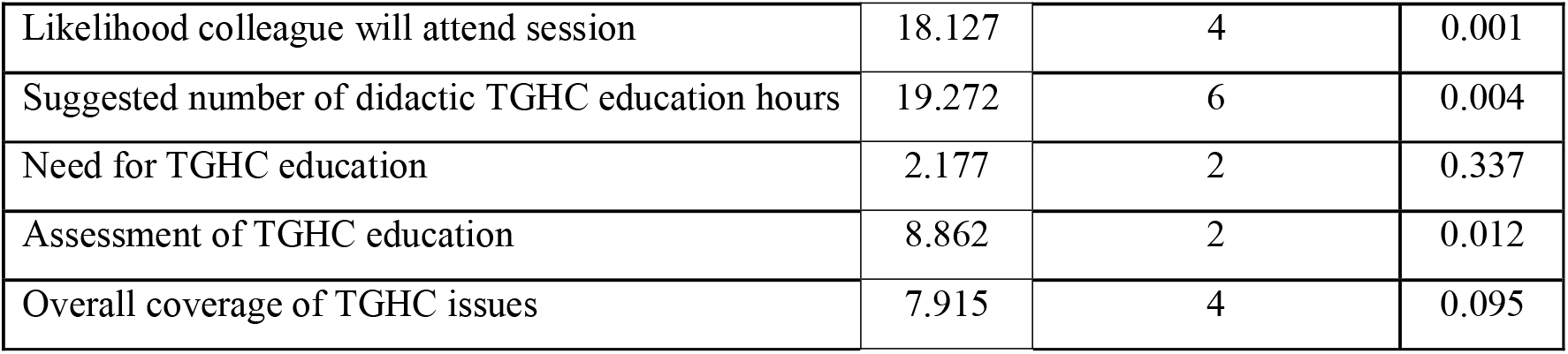
Association of institution region with variables.

| **Table 7. Association of institution region with variables** |  |  |  |
| --- | --- | --- | --- |
| **Variable** | **$\chi^2$ value** | **Degrees of freedom** | **p-value** |
| Transgender patient encounter | 3.661 | 2 | 0.160 |
| Provision of TGHC education | 7.681 | 2 | 0.020 |
| Perceived Importance | 0.572 | 3 | 0.903 |
| Faculty knowledgeable about transgender health | 25.210 | 3 | <0.001 |
| Likelihood participant will attend session | 4.212 | 4 | 0.378 |
| Likelihood colleague will attend session | 18.127 | 4 | 0.001 |
| Suggested number of didactic TGHC education hours | 19.272 | 6 | 0.004 |
| Need for TGHC education | 2.177 | 2 | 0.337 |
| Assessment of TGHC education | 8.862 | 2 | 0.012 |
| Overall coverage of TGHC issues | 7.915 | 4 | 0.095 |

## Discussion

Medical education has been acknowledged as a vital tool for addressing health disparities faced by the transgender population. In order to address the significant healthcare inequality faced by the transgender population in Pakistan, it is vital to address the lack of transgender patient oriented education delivered by most institutions in Pakistan (4). For this purpose, we found it important to scope and understand the perceptions of our medical students regarding the existing transgender healthcare education, as well as the need for an expanded transgender health curriculum. We conducted a cross-sectional study to identify the preferred approaches of medical students in Pakistan on incorporating TGHC education into the curriculum and to identify the possible barriers for incorporating TGHC education into the curriculum.

Out of 436 respondents, 94.7% said that they were not taught any form of transgender healthcare education at their institute. 11 students reported that the annual duration of transgender healthcare education at their institution lasted from 1-3 hours, and a mere 3 respondents reported 7-10 hours of annual transgender education. These findings demonstrate that transgender health remains noticeably absent from the current curriculum, as indicated by the participants’ reports of receiving either no exposure, or very limited hours of exposure to this important concern. This correlates well with a 2020 cross sectional study of a private school in Pakistan, where 69.1% of students surveyed reported poor knowledge of transgender health concerns (4).

In contrast, a study in the US surveying Physician Assistant Programs for transgender health curriculum found that 85.6% of all programs taught transgender health concerns, with 85.6% delivering at least 1-3 hours of content (21). This reveals that programs training healthcare professionals in Pakistan are noticeably deficient and provide significantly lower amounts of transgender health education. This may be linked to the social stigma and taboo status of the transgender population in Pakistan (9), leading to a lack of recognition of the unique health concerns faced by transgender people.

However, it is encouraging to note that all the participating students agreed that it was important to teach healthcare specific to transgender needs at their institution. 98.4% of all participants also agreed on the need for transgender healthcare curriculum. It is a promising sign that medical students recognize the importance of transgender health concerns despite the general lack of awareness in Pakistan. This positive attitude among future doctors could signify a shift in attitudes (22) as well as understanding of the need for specialized and inclusive care. This willingness to learn is an important step suggesting a potential decrease in the healthcare disparity experienced by Pakistan’s transgender population.

To aid our mission to improve transgender health education, we found it important to involve students in the process of developing an engaging and effective curriculum. We asked students about the specific topics they wish to cover about transgender health, as well as their preferred modes of learning and assessment. Our approach is supported by the Adult Learning Theory (23), which suggests the importance of learner participation to ensure that the curriculum is practical and relevant.

Regarding the mode of delivery, students identified current methods at their institutions to include longitudinal themes sessions (10.5%), pre-reading of study materials and didactic lectures (13.2%), and seminars (15.8%). When asked about preferred mode of delivery, problem-based learning (18.1%) and small group discussions (15.9%) were the most prevalent suggestions. It is important to note that students prefer interactive and engaging modes of delivery. As opposed to traditional lectures, active learning has been shown to provide a significant increase in retention and understanding (24). Problem based learning and discussions would encourage the students to apply concepts and think critically which would increase their overall learning.

Among the suggested topics for a future transgender healthcare curriculum, the most prevalent included health access issues/barriers to care faced by transgender persons (14.0%), health disparities faced by transgender persons (13.8%), and mental health among transgender persons (13.8%). A 2022 study in Lahore, Pakistan that investigated transgender health problems and barriers to healthcare found that the most common health problems were depression (56%) and anxiety (59%). A feeling of non-acceptance was a significant barrier to healthcare (25). Therefore, a curriculum that addresses these student suggested topics would be ideal to provide equitable comprehensive care to the transgender community of Pakistan.

The leading perceived barrier to providing transgender health education was transgender health not considered an important issue (17.2%) followed by lack of faculty knowledge about transgender health (13.7%) and lack of time in curriculum (13.2%). This correlates well with a systematic review which identified lack of educational material, lack of expertise and time constraints as common barriers. Moreover, it also identified lack of participation from certain subgroups and costs associated with curriculum development as potential barriers (20). Similarly, a scoping review identified lack of curricular time, lack of competency among faculty and underwhelming institutional support as leading barriers. Therefore, it is important that along with raising awareness about the issue and developing appropriate educational resources an equal emphasis should be placed on training personnel to deliver information regarding transgender health (26).

A major obstacle to incorporating TGHC education was cultural barriers as many participants identified how either the students or faculty were not comfortable discussing the subject (10% and 12.8% respectively). Similarly, many participants identified the culture at the institute not supportive of teaching about transgender health (8.9%). In contrast a study from USA assessing transgender health education in physician assistant programs reported institute’s culture as a barrier in only 7 of the 236 programs evaluated (21). It is rather non-reassuring, although predictable, that many responses included culture to be a barrier to TGHC education. Unlike logistical barriers which might be relatively easy to target, cultural changes take place bit-by-bit over a period making it a tedious process.

We noted that the institutions’ geographical location had a significant association with the provision of TGHC education. This is like a study from USA which reported that the proportion of programs that do and do not teach transgender health varied geographically: 34% of the Program Directors (PDs) who reported that their programs teach this content were in the Northeast, contrasted with 34.7% in the South, 12.4% in the West, and 18.8% in the Midwest. The proportion of programs that teach transgender health additionally varied by program type: 68.8% of programs teaching this content were private institutions whereas 29.7% were public institutions (21). However, there was no significant association between the type of institution (public vs. private) and provision of TGHC education in our study.

Furthermore, there was a significant association between the type of institution and the faculty being knowledgeable about TGHC education. Similarly, there was a significant association between the type of institution and the need for TGHC education. A study conducted in 2018 among endocrinology fellows showed similar results in which majority of participants felt it was important for them to receive training in TGHC education (95.9%). However, only 58.9% reported that TGHC education was a part of their curriculum (27). Our analysis revealed multiple significant associations between the institution region and the variables assessed. This includes a significant association with faculty being knowledgeable about TGHC education and the likelihood of a colleague will attend a session related to the topic. In a similar study, out of 236 physician assistant (PA) programs assessed, 132 (55.9%) programs had at least 1 faculty member knowledgeable in TGHC education (21). One rationale could be the varying literacy rates and acceptance related to TGHC in certain geographical areas as compared to others, thus leading to varying rates of faculty being knowledgeable about the subject. Unfortunately, limited literature exists assessing association between institution type and region with provision of TGHC education and warrants need for more studies determining these associations.

Since the health care and medical education setup of developing countries differ vastly from developed countries, we felt the need to conduct this study in Pakistan. To the best of our knowledge this is the first study from Pakistan assessing student’s views on preferred modalities to receive transgender health education and possible factors that could prove to be barriers in implementation of those measures. The data was collected from medical students across Pakistan giving a good representative sample from the entire country, and thus making the results generalizable to the medical student community in Pakistan. The questionnaire was adapted from a previously validated questionnaire further adding on to the strengths of this study. The questionnaire was limited to 1 per person by using an inbuilt feature in google forms thus making sure that there was no duplication in data.

The limitations of our study include that the different teaching modalities included in our study are adapted from existing literature and might not include other possible interventions. Since we did not carry out any pedagogical intervention ourselves, the feasibility of such interventions in our population remains unclear. Our study focuses solely on the perspectives of medical students and the findings may not be applicable when implementing interventions at other levels such as high schools and during physician training. The questionnaire was circulated online to medical students with the help of peers in other medical colleges so it was not possible to cover every medical college of the country so this could be a reason for a low response rate from institutes in Baluchistan, AJK and KPK and this might influence the generalizability of the study. We believe that our study lays down a solid foundation highlighting the possible modalities which can prove successful in dissemination of transgender health education and can be used as a steppingstone for future studies in which such interventions are implemented, and their efficacy determined. The limitations provide possible opportunities for more research to be conducted on this topic.

## Conclusions and Implications

This study helped to further strengthen the fact that a visible gap exists regarding transgender health education in the medical education system especially in a developing country like Pakistan. The study highlights important barriers and recommends possible modes of delivery to inculcate TGHC education in the curriculum of medical education including incorporation of transgender healthcare education into medical college curricula, and further research among nursing and allied health disciplines. Following on from the lessons we have learned from conducting this study, we present several recommendations. This includes the incorporation of transgender healthcare education into medical college curricula to provide medical students—individuals who are future healthcare professionals—the skills to deal with individuals of all identities and not just the cis-gender status quo. We also recommend the investigation of such surveys in nursing and allied health disciplines as well as other services domains to further minimize the gap in making healthcare and other vital services available to the transgender population. The findings of this study can be extrapolated to the current negligent condition of transgender healthcare coverage across the nation—an observation that provincial governments and their underlying administrations need to improve by introducing policies protecting the interests of transgendered citizens both at the educational level as well as in the healthcare sector.

## Data Availability

All data produced in the present study are available upon reasonable request to the authors.

## Notes

### Competing Interest Statement

The authors have declared no competing interest.

### Funding Statement

This study did not receive any funding.

### Author Declarations

Ethical Review Committee (ERC) at The Aga Khan University Pakistan reviewed this protocol and granted a waiver. ERC number: 2021-6814-19491

